# Predicting Amyloid Positivity Through Proteomic and Machine Learning Approaches

**DOI:** 10.1101/2025.10.20.25337968

**Authors:** Stelios Lamprou, Frank J. Gunn-Moore, Kalliopi Mavromati, Terry J. Quinn

**Author notes:** **SL** is Corresponding Author, School of Cardiovascular and Metabolic Health, University of Glasgow, Glasgow Cardiovascular Research Centre (GCRC), BHF Centre of Research Excellence, 126 University Place, Glasgow, G12 8TA Glasgow, UK.

## Abstract

**Introduction:** Alzheimer’s disease is a progressive neurodegenerative disorder where early detection remains difficult. To address this challenge, we analysed a large proteomics dataset from older adults, including individuals diagnosed through clinical and imaging confirmation of brain amyloid deposition. We hypothesized that amyloid positivity could be detected using blood-based proteomic profiles combined with statistical and machine learning methods.

**Methods:** We applied descriptive and inferential statistical analyses alongside supervised classification approaches, and group comparisons between amyloid positive and amyloid negative individuals were conducted. Classification methods, including random forests, gradient boosting, and neural networks, were used to evaluate prediction of amyloid status. All data were normalized and privacy compliant.

**Results:** Distinct proteomic signatures were associated with disease status. Significant protein expression differences were observed between amyloid positive (*n*=337) and amyloid negative (*n*=651) groups. Classification models reached balanced performance with prediction accuracy up AUC of 0.80. Eight proteins (i.e. SERPINA1, C3, CRP, APOE4, CFH, VTN, C1QTNF5, and PON1) emerged as strong predictors from the best-performing classifiers, representing potential biomarker candidates.:

**Discussion and Conclusions:** Combining statistical and machine learning methods enabled robust identification of patterns distinguishing amyloid profiles. This strategy supports biomarker discovery and development of accessible blood-based diagnostic and therapeutic targets.

**Significance Statement:** This study leverages high-throughput proteomic profiling and machine learning to identify peripheral blood-based protein signatures associated with cerebral amyloid pathology, a hallmark of Alzheimer’s disease. By integrating clinical data with proteomic biomarkers, we aimed to develop a non-invasive, scalable predictive tool that can support early detection and risk stratification of Alzheimer’s disease, potentially improving screening efficiency and guiding future therapeutic strategies. Furthermore, this analysis allowed for mechanistic insight into the biology of amyloid in Alzheimer’s disease.

**Highlights:**

- Developed a machine learning-based proteomic model to predict amyloid positivity in Alzheimer’s disease using plasma blood samples.
- Identified a protein machine learning generated signature linked to central amyloid pathology.
- Suggested the potential for scalable, early detection tools to support precise and targeted diagnosis and treatment planning in Alzheimer’s disease using machine learning.

## Introduction

Alzheimer’s disease (AD) is a chronic progressive neurodegenerative disorder, typically beginning with memory loss. As the condition progresses, it can impact behaviour, language, spatial awareness, and motor functions [1]. It is also the leading cause of dementia [2], with an escalating global burden that demands novel strategies for earlier and more accurate diagnosis [3].

A critical pathological hallmark of AD is the accumulation of amyloid-beta (Aβ) plaques in the brain, which can be detected through positron emission tomography (PET) scans or cerebrospinal fluid (CSF) analysis [4–6]. However, these methods are either invasive, expensive, or not widely accessible, posing limitations for routine screening, population-level monitoring, or early detection efforts [7, 8]. In response to this diagnostic gap, there is increasing interest in identifying peripheral biomarkers, particularly blood-based signatures, that reflect central amyloid pathology [9, 10].

Plasma proteomics, the high-throughput measurement of protein expression in blood samples, offers a minimally invasive approach to uncover systemic biological signatures associated with AD and brain ageing [11–13]. When combined with advanced analytical tools such as machine and deep learning (ML/DL), proteomic data can be harnessed not only to discover novel biomarkers but also to build predictive models capable of classifying disease and healthy states [14–16]. ML/DL algorithms such as random forests, gradient boosting machines, and neural networks are particularly well-suited for this task due to their ability to handle large feature spaces and complex interactions between variables [17, 18].

Supervised ML are computational algorithms designed to learn patterns from labeled data, enabling them to categorize new, unseen instances into predefined classes or predict continuous outcomes. These models operate by mapping input features to target variables through a training phase, during which the algorithm iteratively refines its parameters to minimize prediction error. DL, a specialized subset of machine learning, utilizes artificial neural networks with multiple layers to capture complex, non-linear relationships within high-dimensional data. Collectively, artificial intelligence encompasses these methodologies to simulate human-like decision-making and problem-solving abilities. In the context of supervised classification for predictive modeling, these models are invaluable for their ability to improve decision-making processes by accurately assigning class labels based on input data characteristics. Performance evaluation involves assessing discrimination ability, often quantified via metrics such as the area under the receiver operating characteristic curve (AUC), alongside calibration of predicted probabilities to ensure reliability. To avoid overfitting, where a model performs well on training data yet poorly generalizes to new data, techniques such as cross-validation and hyperparameter tuning are employed. Despite inherent model imperfections, supervised ML models offer powerful tools for reliable prediction and classification tasks across diverse applications in biomedical and clinical research [18, 19].

In this study, we analyzed a rich dataset comprising plasma proteomic measurements from people living with Alzheimer’s disease by employing such ML/DL models. Our primary objective was to assess the ability of proteomic profiles to distinguish amyloid status, as determined by PET scan outputs and not to distinguish clinical AD from ‘normal’. The rationale behind this classification task stems from the hypothesis that specific proteins, alone or in combination, may serve as peripheral indicators of central amyloid pathology.

Our specific aims were: (i) To perform classical statistical analyses to explore differences in protein expression across the amyloid subgroups and identify candidate biomarkers associated with amyloid status. (ii) To use these datasets to inform subsequent ML analyses, build predictive models for amyloid classification, and validate the models. (iii) To compare feature importance across models to identify a reproducible core proteomic signature and then assess potential biological relevance to dementia.

## Materials and Methods

### Study Objective and Design

This study aimed to assess whether plasma proteomic profiles can effectively distinguish between individuals with positive and negative amyloid status. The overall analytical pipeline combined classical statistical methods with supervised ML/DL approaches. The goal was twofold: (i) to identify proteomic features that differentiate amyloid-positive and amyloid-negative subjects and (ii) to evaluate the predictive power of ML/DL classifiers trained on these features.

### Data Source

This research drew data from the Bio-Hermes Study, a comprehensive, multi-center project designed to build a database of blood-based and digital biomarkers for AD among community-dwelling individuals from diverse backgrounds (Mohs et al., 2024). PET imaging with ^18F-Florbetapir was utilized to assess cerebral amyloid burden. Amyloid status was determined using a standardized uptake ratio (SUVR) cutoff of 1.1, corresponding to a Centiloid scale value of 24.1, which aligns with thresholds employed in contemporary Alzheimer’s disease clinical trials. Notably, 21% of participants who were cognitively normal exhibited amyloid positivity, consistent with preclinical AD pathology. Between April 2021 and November 2022, a total of 1,296 participants were enrolled across 17 research sites in the United States, with 1,001 ultimately included in the final dataset. To ensure representation, the Global Alzheimer’s Platform implemented targeted recruitment efforts that successfully increased participation from under-represented groups, resulting in 24% of the sample identifying as minorities. Full eligibility and exclusion criteria have been described in Bio-Hermes primary papers (Mohs et al., 2024).

### Data Preparation and Preprocessing

Plasma proteomic measurements were collected from individuals with known amyloid status by EMTherapro (https://emtherapro.com/, Georgia, USA) from a sub-cohort (*n*=1001). Amyloid status was identified using PET scans and proteomics measurements were conducted using tandem mass spectrometry (TMTPro18 pipeline, Thermo Fisher Scientific). The final dataset provided included 992 participants after quality control and normalization (i.e. abundance of the protein was normalized by overall total protein). The total proteins were 881 (Supplementary (Sup.) Table 1). Prior to analysis, all protein features, and subject samples with missing values, were excluded to ensure clean input for downstream modelling resulting in a dataset of 295 proteins and 988 subjects as seen in Figure 1. From these 295 proteins 31 that were found to be differentially expressed between the amyloid status groups were further selected as features for ML training. The final sample size for each class was 337 for positive amyloid and 651 for negative amyloid.

**Figure 1.**
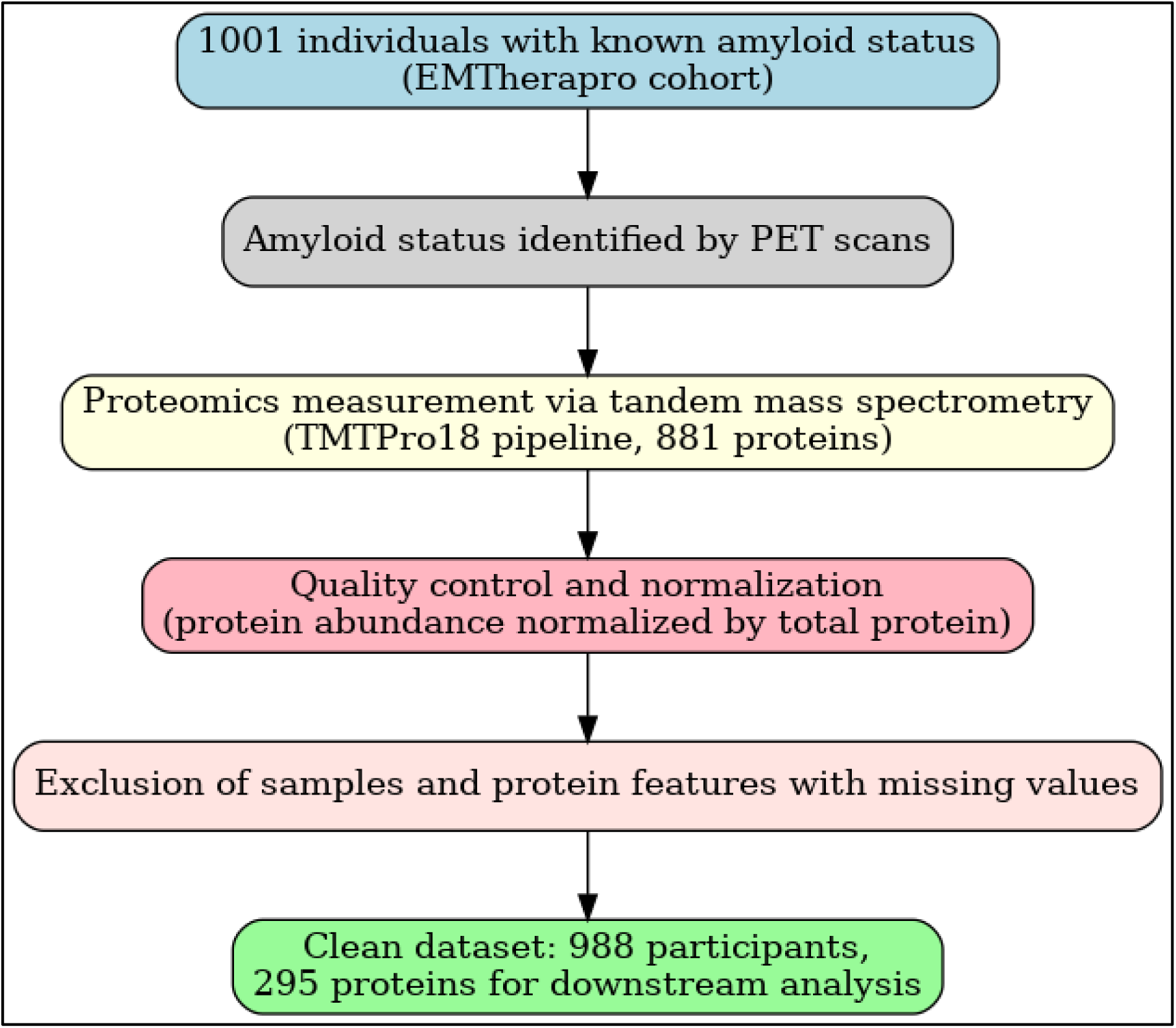
Data Preparation and Sample Size. Generated using Perplexity

**Table 1.** Demographics of Patient Cohort – Bio-Hermes/EMTherapro Characteristic.

| Characteristic | Value |
| --- | --- |
| Total sample size | 988 |
| Age (years) | Range: 59 – 85<br>Mean: 72<br>Median: 72 |
| Sex % (*n*) | Female: 56% (553)<br>Male: 44% (435) |
| Race/Ethnicity<br>% (*n*) | White: 85.6 (845)<br>Black or African American: 11.5 (113)<br>Asian: 1.9 (19)<br>American or Alaska Native: 0.2 (2)<br>Native Hawaiian or Other Pacific Islander: 0.1 (1)<br>Unknown: 0.8 (8) |
| Amyloid Status<br>%(*n*) | Positive: 34 (337)<br>Negative:66 (651) |

### Feature Selection

To shortlist features for model training, classical statistical methods were applied as an initial screening step. Two-sample independent t-tests were conducted (t-test assumptions were tested prior to running these tests, and all met these requirements) to compare mean protein levels between amyloid-positive and amyloid-negative groups. The proteins with significant differential expression (i.e. FDR <0.05) between groups were chosen as features to train the ML models. FDR correction resulted in five differentially expressed proteins. As this number was low as a basis for ML training, we expanded the protein set by included those showing between group differences prior to correction for multiplicity (i.e. unadjusted p-value <0.05). This resulted to 31 chosen proteins out of 295.

### Machine Learning Pipeline

Three supervised machine learning algorithms were employed to evaluate predictive performance: Random Forests (RF), Gradient Boosting (GB), and Neural Networks (NN). Each model was trained and tested across four distinct training/testing splits (60/40, 70/30, 80/20, and 90/10), employing a fixed random seed (set.seed(123)) to ensure reproducibility. Class imbalance was observed and addressed using the ‘downSample’ function, which randomly downsizes the majority class equal to the minority class sample size; each class ended up with the same sample size (*n*=337) which reflected the sample size of the minority class (i.e. positive amyloid).

Hyperparameter tuning and repeated cross-validation were conducted within each algorithm to optimize predictive accuracy and mitigate overfitting. The caret package in R facilitated these procedures, where the ‘tuneLength’ parameter guided the number of different hyperparameter values evaluated during model training. Stability and generalizability of the models were assessed via 10-fold repeated cross-validation, repeated 100 times to yield 100 Random Repeats (RR).

For each model, confusion matrices and Receiver Operating Characteristic (ROC) curves were generated to compute performance metrics including Area Under the Curve (AUC), Sensitivity, Specificity, Balanced Accuracy (BA), and Positive/Negative Predictive Values (PPV/NPV). Feature importance rankings were extracted from each model to elucidate predictor contributions. To compare classification performances, ROC-AUC was employed as the discrimination metric, and DeLong’s test for correlated ROC curves, adjusted with the Benjamini-Hochberg procedure for multiple comparisons, was applied to determine statistically significant differences between models.

Data preprocessing, manipulation, analysis, and principal visualizations were executed using R version 4.5. Core analytical workflows leveraged the caret package, which enabled streamlined model training, hyperparameter tuning using ‘tuneLength’, and performance assessment through repeated cross-validation. Data handling and graphical representations were supported by the tidyverse suite, comprising dplyr and ggplot2. Additional machine learning frameworks were implemented with ranger for Random Forests, gbm for Gradient Boosting, and nnet for Neural Networks.

### Feature Integration and Comparative Analysis

To identify consistently predictive features, the top 10 proteins from each of the best-performing RF, GB, and NN models were compared. The analysis focused on the first 10 features, as the importance scores for subsequent variables showed a steep decline, with lower-ranked features contributing minimal predictive value. In the caret package in R, RF determines feature importance through the mean decrease in Gini impurity or classification accuracy, while GB models assess importance by summing the improvements in the loss function attributed to each feature across all trees. For NN, variable importance is typically derived by analyzing the connection weights between input and hidden layers, reflecting the relative influence of each input on the model’s predictions. Overlapping features across models were visualized using Venn diagrams. Proteins that appeared in all three top-performing models of each learning method or were shared between the most accurate (RF and GB) were selected as part of a consensus proteomic signature.

### Pathway and Disease Association Analysis Using IPA

To explore the biological relevance of the shortlisted proteins identified through machine learning, we employed the Pathway Analysis function of Ingenuity Pathway Analysis (IPA, Qiagen, Manchester, UK). The analysis was conducted under default settings, with species set to *Homo sapiens* and all tissues and cell lines included. Using the Connect-Pathway function, we examined known molecular interactions, canonical pathways, and disease-associated networks enriched in our protein set.

IPA’s Pathways and Diseases & Functions modules were used to assess whether any of the shortlisted proteins had documented associations with Alzheimer’s disease, dementia, or broader neurodegenerative processes. The resulting networks and annotations were filtered for neurological disease relevance, and high-confidence (experimentally observed) relationships were prioritized. Enrichment scores were calculated using Fisher’s exact test, and pathway/disease associations with p-values < 0.05 were considered significant. Key findings, including any direct or indirect involvement of proteins in amyloid processing, tau phosphorylation, neuroinflammation, or synaptic function, were further evaluated for biological plausibility based on published literature.

### Data and Code Availability

Data of the Bio-Hermes Data Challenge study are publicly available and hosted at the Alzheimer’s Disease Data Initiative Platform (https://www.alzheimersdata.org/) through the **AD Discovery Portal on AD Workbench,** while the general code for ML pipeline can be found on GitHub (https://github.com/stelioslamprou37/SML_PipelineR).

### Ethics and Reproducibility

All analyses were conducted using standardized and transparent workflows. Model training and testing procedures were fully reproducible due to the use of fixed seeds and documented data preprocessing steps. As this study relied on secondary, de-identified data, it was conducted in compliance with all relevant ethical standards. At time of analysis, data were only available through the Alzheimer’s Disease Data Initiative (alzheimersdata.org) initially for researchers participating in the Bio-Hermes Data Challenge [20]. Data are now open source at this platform.

## Results

### Patient Characteristics and Demographics

### Sample Overview and Feature Selection

Following initial data curation, the dataset comprised 988 samples and 295 proteins. Five proteins were found to have significant differential expression accounting for multiple analyses: ApoE4 (FDR=0.0000000000002299), P01024 (FDR=0.00075117), P08603 (FDR=0.023618), P02671 (FDR=0.0340305), and P04004 (FDR=0.0341628). As this was considered a low feature size for predictive modelling proteins with unadjusted p-value < 0.05 were also considered. That led, in a total of 31 proteins were found to be differentially expressed significantly development (Sup. Table 3).

### Machine Learning Analysis

Model performance can be found in Table 2. Moreover, a total of 66 DeLong’s test comparisons were conducted across three model families: RF (models 1–4), GB (models 5–8), and NN (models 9–12). The resulting adjusted p-values are visualized in a heatmap (Figure 2), with colour intensity reflecting the degree of statistical significance.

**Figure 2.**
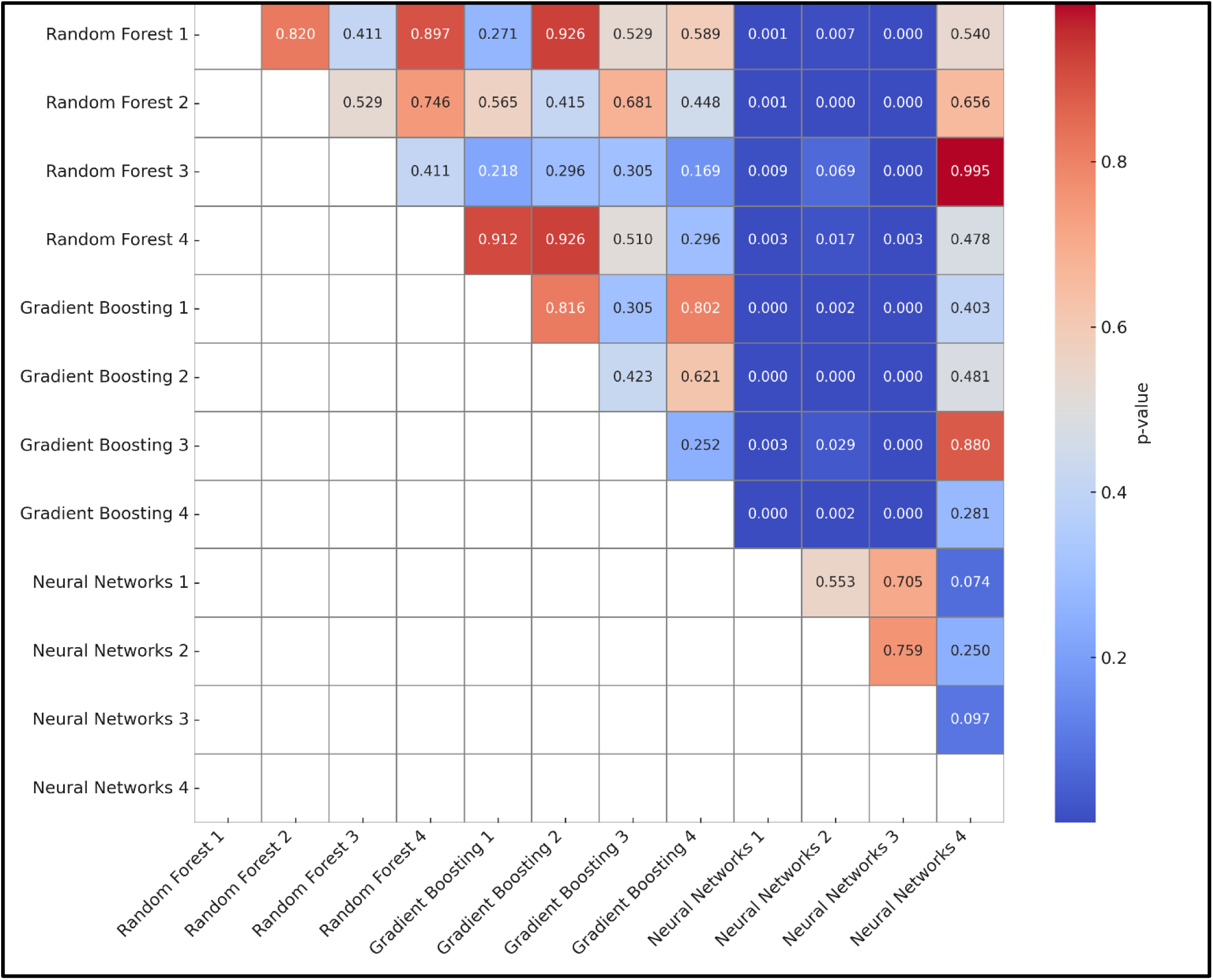
Pairwise Comparison of Model Performance Using DeLong Test p-values. This heatmap visualizes pairwise statistical comparisons between classification models based on DeLong test p-values (BH-corrected) for ROC-AUC. Lower p-values (closer to 0, shown in blue) indicate statistically significant differences in performance between the models, while higher p-values (closer to 1, shown in red) suggest no significant difference. Only the lower triangle of the matrix is populated due to symmetry in comparisons. *Generated using R v.4.5*

**Table 2.** ML Performance Metrics.

| Metric | RF1 | RF2 | RF3 | RF4 | GB1 | GB2 | GB3 | GB4 | NN1 | NN2 | NN3 | NN4 |
| --- | --- | --- | --- | --- | --- | --- | --- | --- | --- | --- | --- | --- |
| **AUC** | 0.8 | 0.7 | 0.7 | 0.8 | 0.8 | 0.8 | 0.7 | 0.8 | 0.5 | 0.6 | 0.6 | 0.7 |
| **NPV** | 0.8 | 0.7 | 0.6 | 0.7 | 0.8 | 0.7 | 0.6 | 0.7 | 0.6 | 0.6 | 0.6 | 0.6 |
| **PPV** | 0.6 | 0.7 | 0.7 | 0.7 | 0.6 | 0.7 | 0.6 | 0.7 | 0.5 | 0.5 | 0.5 | 0.7 |
| **BA** | 0.7 | 0.7 | 0.6 | 0.7 | 0.7 | 0.7 | 0.6 | 0.7 | 0.5 | 0.6 | 0.6 | 0.7 |
| **Specificity** | 0.6 | 0.7 | 0.7 | 0.6 | 0.6 | 0.7 | 0.7 | 0.7 | 0.5 | 0.5 | 0.3 | 0.7 |
| **Sensitivity** | 0.8 | 0.7 | 0.6 | 0.8 | 0.8 | 0.7 | 0.6 | 0.7 | 0.6 | 0.6 | 0.8 | 0.6 |
| **Accuracy** | 0.7 | 0.7 | 0.6 | 0.7 | 0.7 | 0.7 | 0.6 | 0.7 | 0.5 | 0.6 | 0.6 | 0.7 |

\*BA: Balanced Accuracy / NPV-PPV: Negative-Positive Predictive Value / AUC: Area Under the Curve

RF and GB yielded the highest and most consistent classification performance metrics across all splits (i.e. DeLong’s test output indicated no significant difference between these models.). AUC values ranged from approximately 0.7 to 0.8, with the strongest performance observed in the 80/20 and 90/10 training/testing splits (AUC=0.8) for both methods.

The NN model exhibited AUC values between 0.5 and 0.7 across splits. This demonstrates less performance than the RF and GB counterparts (i.e. AUC: 0.7-0.8), which is also supported by significant differences when compared using DeLong’s tests.

To identify consistently important proteins, we examined the top 10 ranked features from the best performing model of each learning method. Features were extracted (Sup. Table 2) and compared. This threshold was chosen pragmatically to focus on the most influential predictors while limiting noise from lower-ranked variables, and to facilitate a clear and interpretable comparison across models. Feature importance for RF and GB was calculated using mean decrease in Gini and gain scores, respectively. For NN, feature weights from the input layer were used to approximate relative influence. Based on the overall metrics obtained the 90/10 training/testing split models for all methods (i.e. RF-4, GB-4 and NN-4) achieved an efficient level of performance with BA around 70% and AUC ranging from 70-80%. From these 3 models, the top 10 most important features were extracted and compared.

Based on these comparisons (see Figure 3), 4 proteins were ranked as top 10 important features from all classifiers, these are P04004, P08603, Q13740, and P01024. We also assessed the overlap between RF-4-GB-4 alone, as these 2 models performed in a very efficient manner, surpassing the performance of NN-4. For this comparison the following proteins were overlapped: ApoE4, P01009, P02741, andQ6ZS72. Table 3 summarises these 8 proteins.

**Figure 3.**
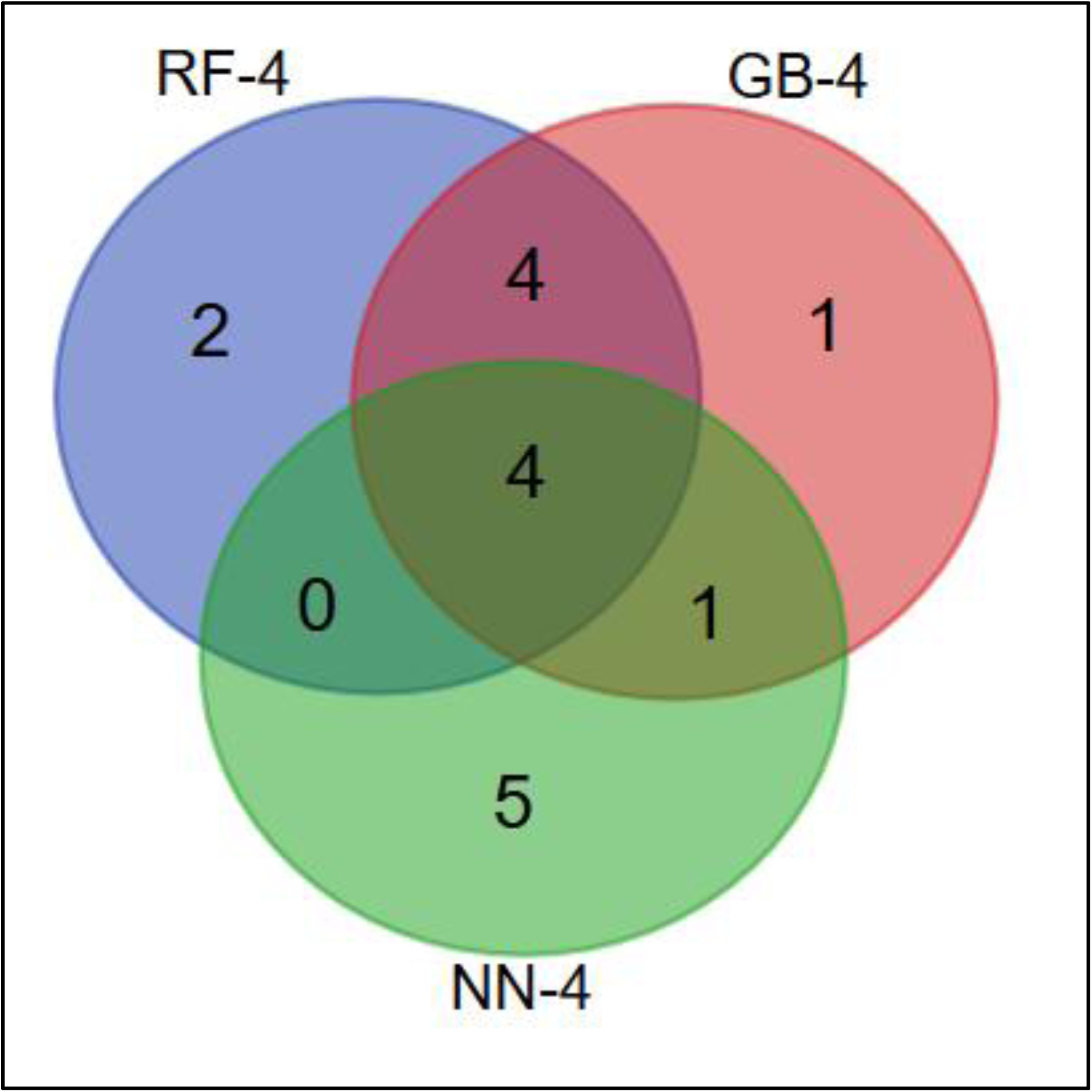
Overlap of Top-10 Proteins Across Machine Learning Models: RF-4, GB-4, NN-4. The Venn diagram illustrates the overlap of the top-ranked protein features identified by three machine learning models: RF (RF-4; blue), GB (GB-4; red), and NN (NN-4; green). A core set of four proteins (P04004, P08603, Q13740, and P01024) was shared across all three models, indicating strong agreement on their importance for classification. An additional four proteins (P01009, P02741, ApoE4, and Q6ZS72) were shared between GB-4 and RF-4, highlighting a high level of concordance between the two ensemble-based methods. GB-4 and NN-4 shared one protein (P01008), while RF-4 uniquely identified two proteins (P02748 and P00742), and GB-4 identified one unique protein (P02750). The neural network (NN-4) selected five unique proteins (P01042, P04114, Q12805, P05090, and Q15113). *Generated using* https://bioinformatics.psb.ugent.be/webtools/Venn/

**Table 3.** Proteins with Predictive Ability of Amyloid Positivity.

| UniProt ID* | Protein Name | Gene Symbol | Function |
| --- | --- | --- | --- |
| **P04004** | Vitronectin | **VTN** | Involved in cell adhesion, coagulation, and inhibition of the membrane attack complex. |
| **P08603** | Complement factor H | **CFH** | Regulates the complement system to prevent damage to host cells. |
| **Q13740** | Serum paraoxonase/aryl-esterase 1 | **PON1** | Detoxifies organophosphates and protects lipoproteins from oxidative damage. |
| **P01024** | Complement C3 | **C3** | Central component of the complement cascade; marks pathogens for destruction. |
| **P01009** | Alpha-1-antitrypsin | **SERPINA1** | Protease inhibitor that protects tissues from enzymes of inflammatory cells. |
| **P02741** | C-reactive protein | **CRP** | Acute-phase protein that binds to phosphocholine on dying cells and pathogens. |
| **ApoE4** | Apolipoprotein E, Isoform E4 variant | **APOE** | Lipid transport and injury repair in the CNS; E4 isoform linked to Alzheimer's risk. |
| **Q6ZS72** | Complement C1q tumour necrosis factor-related protein 5 | **C1QTNF5** | Involved in immune response and possibly retinal structure maintenance. |

\*Group-1: Negative Amyloid and Group-2: Positive Amyloid. A positive *t*-value (blue) indicates that the protein is upregulated in the Amyloid-negative group, while a negative *t*-value (red) suggests upregulation in the Amyloid-positive group.

Among the eight proteins of interest, four (i.e. **P01024**, **P02741**, **P08603**, and **Q13740**) appear to be upregulated in Amyloid-positive individuals (see Sup. Table 3). Conversely, the remaining four proteins (i.e. **P04004**, **Q6ZS72**, **P01009**, and **P05090**) exhibit upregulation in the Amyloid-negative group.

### Protein Association with Dementia and AD

IPA outputs of reported-in-literature associations with AD and construct such pathway are illustrated in Figure 4. Based on reported findings, upregulation of the human SERPINA1 protein in cerebrospinal fluid and increased expression of the human C3 protein in cortical brain tissue are both associated with AD in humans [21, 22].

**Figure 4.**
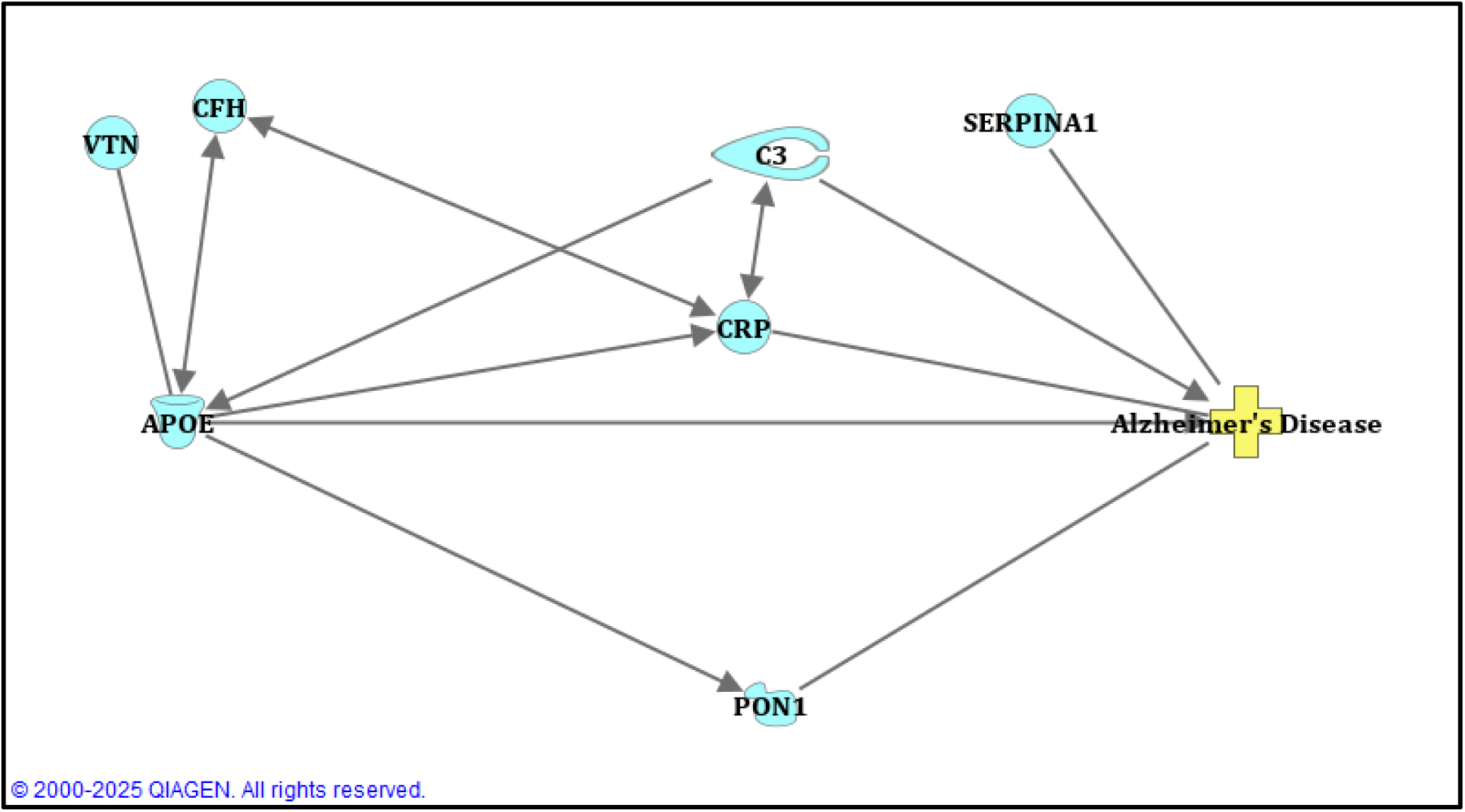
Key Protein Associations with Alzheimer’s Disease Pathology. Upregulation of SERPINA1 in cerebrospinal fluid and increased C3 expression in cortical tissue are linked to Alzheimer’s disease (AD). Elevated plasma CRP, which activates C3B, also associates with AD pathology. The APOE4 protein and the rs429358 SNP mutation in the *APOE* gene influence CRP localization and mediate interactions with complement factor H (CFH) and vitronectin (VTN). In mouse models, Apoe knockout decreases PON1 activity, while mutation of the C3 gene increases Apoe mRNA in the retina. A mutant human PON1 variant (p.Q192R) is implicated in AD, highlighting its role in disease mechanisms. *\*Rows indicate direct interactions (e.g., one molecule promotes another’s expression), while no arrow reflects associations, such as APOE variants linked to higher AD risk.**Different shapes indicate a different type of molecule or state; Diseases are represented as a **Cross**. APOE is represented by a **transporter protein icon**. Group of proteins or complexes are illustrated by a **Circle**. PON1 icon represents a **Phosphatase.** Generated using IPA-QIAGEN*

Additionally, elevated levels of human CRP protein in plasma, which promotes activation of the C3B protein [23], are linked to AD pathology [24]. The human APOE4 protein plays a significant role in AD incidence [25, 26], and mutation of the human *APOE* gene (single nuclear polymorphism substitution rs429358) influences the localisation of CRP in serum [27], as well as mediates binding interactions between APOE, complement factor H [28], and vitronectin [29]. For CFH and VTN, IPA was unable to detect any reported associations.

In mouse models, knockout of the *Apoe* gene reduces the activity of the PON1 protein [30], while mutation of the *C3* gene in the rd10 mouse retina leads to increased Apoe mRNA expression [31]. Furthermore, a mutant form of human PON1 protein (p.Q192R) has been implicated in AD, underscoring the involvement of this protein in disease mechanisms [32]. No association was reported for C1QTNF5.

## Discussion

This study demonstrates that plasma proteomic profiling, when combined with ML, can effectively classify amyloid status in individuals living with AD. Notably, our models achieved strong predictive performance using proteomic data alone—without incorporating clinical or imaging measures—highlighting the promise of blood-based biomarker approaches. Furthermore, certain proteins that comprised these signatures were bioinformatically analysed leading to identification of already reported disease related associations but also one protein (C1QTNF5) with no association These differential expression patterns highlight the potential utility of these proteins in distinguishing amyloid status and suggest mechanistic links worth further investigation.

### ML Model Performance on Predicting Amyloid Positivity

Among the tested algorithms, decision tree-based models (i.e. RF and GB) consistently outperformed NN across all performance metrics. Among them, RF-4 and GB-4 emerged as the top models both achieving high AUC, highlighting their robustness across different training-test splits. NN, in contrast, demonstrated greater variability and generally lower predictive reliability. These findings suggest that ensemble tree-based methods are better suited for proteomics-based classification in dementia research, offering more stable and interpretable results for identifying cognitively healthy individuals. This was also observed in a similar study of assessing the prognostic potential of proteomic profiles for AD using similar methodology like RF models [33].

Although NN performance was poorer than RF and GB, these metrics remained within acceptable classification margins. The somewhat reduced performance of NN may be attributed to its higher sensitivity to sample size and the need for more extensive tuning. A comparative machine-learning study using ADNI data demonstrated that RF maintained accuracy more consistently than NN when feature sets were reduced, indicating that neural networks can be more sensitive to smaller sample sizes and hyperparameter tuning—with greater variability in performance [34]. However, NN still identified several key features that overlapped with those selected by RF and GB, offering complementary validation of the identified signal.

Based on this outputs, we suggest that ensemble learning methods appear particularly suited for handling the complexity of high-dimensional proteomic data, a finding in line with previous works in omics-based ML classification that found that ensemble tree methods like RF and GB offer greater stability and interpretability in high-dimensional, small-sample contexts, while NN still recover overlapping key features—providing complementary validation for biomarkers identified by tree-based models [35].

### Proteins as Predictors of Amyloid Positivity

Crucially, a core proteomic signature of eight proteins emerged as consistently top-ranked features across RF and GB. These proteins may serve not only as classification features, but also as biologically meaningful markers linked to AD pathology. Based on IPA analysis, the seven of eight candidate proteins (i.e. SERPINA1, C3, CRP, APOE4, CFH, VTN, PON1) showed potential relevance to Alzheimer’s disease mechanisms through documented perturbations in human and/or mouse models.

#### Complement Factor H (CFH) & Vitronectin (VTN)

IPA did not reveal human AD literature linking CFH or VTN directly to disease, but APOE4’s **impaired interaction with CFH** reduces FH-mediated regulation of complement in amyloid clearance, offering an indirect mechanistic connection [36]. Vitronectin emerges as a multifaceted player in AD: it has been shown to colocalize with amyloid beta peptide-containing plaques and neurofibrillary tangles in Alzheimer’s disease, underscoring its spatial and structural association with hallmark AD pathology [37].

Moreover, it accumulates within plaques and possesses inherent amyloidogenic capacity yet also exhibits anti-aggregation effects on Aβ in vitro. This paradox suggests that VTN might initially act to sequester and stabilize Aβ oligomers, but aberrant misfolding or overaccumulation within the extracellular matrix could ultimately contribute to plaque consolidation and neurotoxicity [38, 39].

#### α₁-antitrypsin & complement C3

Several studies report elevated specific cerebrospinal fluid SerpinA1 levels in AD, along with disease-specific isoform patterns that correlate with lower Aβ42/40 ratios and increased p-tau/t-tau levels in specific cerebrospinal fluid [36, 40]. Deposits of α₁-antitrypsin have also been identified within Aβ and tau aggregates in post-mortem AD brain, suggesting involvement in protein misfolding and neuroinflammation [40].

Human AD brains exhibit increased C3 expression, particularly co-localizing with synapses, and elevated C3 fragments are present in AD CSF in proportion to tau [41]. In APP and tauopathy mouse models, genetic deletion of C3 reduced synapse loss, neurodegeneration, and improved behaviour—showing C3 contributes to Aβ- and tau-driven pathology [41].

#### C-reactive protein

High plasma CRP is linked to accelerated cognitive decline and increased tau markers (p-tau and t-tau) in APOE ε4 homozygous individuals, indicating CRP may mediate APOE4-associated neurodegeneration [42]. However, mendelian and cohort studies also show lifelong low CRP may paradoxically associate with higher dementia risk [43] that highlights a complex, context-sensitive roles in inflammation and amyloid clearance.

#### APOE4

The ε4 allele remains the single strongest genetic risk factor for late-onset AD. APOE4 is less effective at promoting amyloid-β clearance, contributing to its accumulation [44]. Moreover, APOE4 shows reduced binding to complement factor H, impairing formation of factor H/APOE/Aβ complexes that normally limit Aβ oligomerization and inflammatory toxicity [36]. APOE4 also contributes to blood-brain barrier breakdown, independently of amyloid and tau pathology, accelerating cognitive decline [45].

#### Paraoxonase 1 & C1QTNF5

The PON1 Q192R and L55M polymorphisms modulate serum PON1 activity, which influences oxidative stress and lipid metabolism pertinent to AD. The R allele (Q192R) and M55M genotype are associated with altered brain Aβ levels, cholinergic dysfunction, and increased AD risk [46]. Low PON1 enzymatic activity correlates with lower extracellular Aβ42/Aβ40 ratios, supporting a role in Aβ scavenging and oxidative protection [47]. Moreover, PON1, has been found to be crucial for counteracting oxidative stress, and its decreased activity may contribute to systemic and cerebral oxidative damage and impaired cholesterol metabolism, factors relevant to AD progress. Epidemiological studies show conflicting results regarding PON1 gene polymorphisms and AD risk, but low PON1 aryl-esterase activity is frequently associated with AD [48].

To date, no peer-reviewed association has been reported between C1QTNF5 and AD or amyloid pathology. IPA did not identify relevant literature, and broader searches confirm the absence of documented links. However, integrated analyses of DNA methylation (5 mC, 5hmC) and gene expression in AD revealed that the AKT3 and MBP genes are consistently downregulated in AD patients and mouse models, with significant promoter methylation changes at key CpG sites. The study found 13 promoter CpG sites in genes including C1QTNF5 and AKT3 with differential methylation and expression, correlating these epigenetic alterations with AD pathology. Functional validation in an AD mouse model showed reduced AKT3 expression in cortex and hippocampus linked to cognitive deficits and identified ten chemicals as potential new therapeutic candidates. The results highlight AKT3 and MBP as promising biomarkers and suggest that integrated epigenetic profiling can accelerate novel target identification for AD [49].

### Strengths and Limitations

Key strengths of our analytical approach include rigorous preprocessing, the use of fixed random seeds, multiple supervised algorithms, and a range of data partitioning strategies (e.g., 60/40 to 90/10 splits). This multi-pronged approach enhanced reproducibility and robustness. However, limitations remain. Our models were trained and tested only on proteomic data; in clinical practice, integrating multimodal biomarkers (e.g., imaging, cognitive scores, genomics) could further improve discrimination. Moreover, while we identified proteins strongly associated with amyloid status, our analysis does not establish causality or any more informative biological insight about any association with AD pathophysiology. However, it opens the opportunity of identifying new potential biochemical pathways involved in the pathogenesis of the disease. Finally, the shortlisted proteins were predominantly identified as significantly differentially expressed based on uncorrected p-values, with only five proteins remaining significant after FDR correction (FDR < 0.05), indicating an increased risk of Type I error; however, a less stringent cutoff was necessary to retain a sufficient number of features for effective training of classification models, as overly stringent correction can lead to exclusion of potentially informative predictors and impair model performance by reducing the feature space required to capture the underlying biological variability.

### Clinical and Research Implications

From a translational perspective, the ability to stratify amyloid status non-invasively using plasma proteins could support earlier, scalable AD screening [50–52]. However, for clinical implementation, these proteomic markers would likely need to be combined with other biomarker modalities to optimize accuracy [53]. More intriguingly, the recurrent identification of certain proteins suggests potential new targets for mechanistic study and therapeutic intervention [54, 55]. Further research should investigate these proteins’ roles in AD pathophysiology, validate them in external cohorts, and explore their utility in predicting disease progression or treatment response.

### Future Directions

While our results are promising, several important next steps remain. First, external validation in independent cohorts is essential to confirm the reproducibility and generalizability of the core proteomic signature. Second, the question of causality remains open. Although this study did not include mendelian randomisation analysis, the consistent emergence of certain proteins (e.g. vitronectin) as predictive features supports their potential relevance in AD pathophysiology. Future hypothesis-driven MR studies using large biobanks such as UK Biobank may help to clarify whether these markers are simply correlates of disease or play a causal role in amyloid accumulation.

Third, integrating proteomic data with other modalities (e.g. imaging, multi-omics, and cognitive performance data) could enhance prediction accuracy and offer a more holistic view of AD progression. Multimodal models may also better capture disease heterogeneity, an important consideration for clinical translation. Finally, while this study focused on classification, longitudinal data could support predictive modelling of disease progression. Investigating whether proteomic changes precede amyloid conversion or cognitive decline would provide valuable insights into early disease mechanisms.

## Conclusion

This work presents a reproducible and well-validated framework for using plasma proteomic data and ML to predict amyloid status in AD. The identification of a shared proteomic signature across top-performing models underscores the potential utility of blood-based biomarkers for non-invasive diagnosis and disease monitoring. Our results not only validate the utility of known markers such as ApoE4 but also highlight emerging proteins with strong predictive ability for amyloid positivity like C1QTNF5 as potentially relevant to AD. Although we do not yet test causality, the consistency of these findings suggests a meaningful relationship that warrants further exploration, potentially through future mendelian randomization studies. While further validation and causal investigation are warranted, these findings contribute a solid foundation for future biomarker discovery and translational research in AD.

## Data Availability

All data produced are available online at https://www.alzheimersdata.org/ad-workbench

## Author Contributions (CRediT)

**S. Lamprou:** Conceptualization, Data curation, Methodology, Software, Formal analysis, Visualization, Writing; original draft, Writing; review & editing

**K. Mavromati:** Data curation, Writing; review & editing

**F.J. Gunn-Moore:** Writing; review & editing

**T.J. Quinn:** Conceptualization, Writing; review & editing, Supervision

## Abbreviations

AD: Alzheimer’s disease
Aβ: Amyloid-Beta
APP: Amyloid Precursor Protein
APOE4: Apolipoprotein E4
AUC: Area Under the Curve
BA: Balanced Accuracy
BH: Benjamini-Hochberg
CSF: Cerebrospinal Fluid
DL: Deep Learning
FDR: False Discovery Rate
GB: Gradient Boosting
IPA: Ingenuity Pathway Analysis
ML: Machine Learning
NPV: Negative Predictive Value
NN: Neural Networks
PSV: Positive Predictive Value
PET: Positron Emission Tomography
RF: Random Forest
RR: Random Repeat
ROC: Receiver Operating Characteristic

## Acknowledgments

We gratefully acknowledge the Global Alzheimer’s Platform and the Alzheimer’s Disease Data Initiative (ADDI) for providing access to data and cohorts through the Bio-Hermes Data Challenge. We also thank our colleagues and peers whose valuable contributions and collaboration greatly supported this study.

---

**Sup. Table 1.**
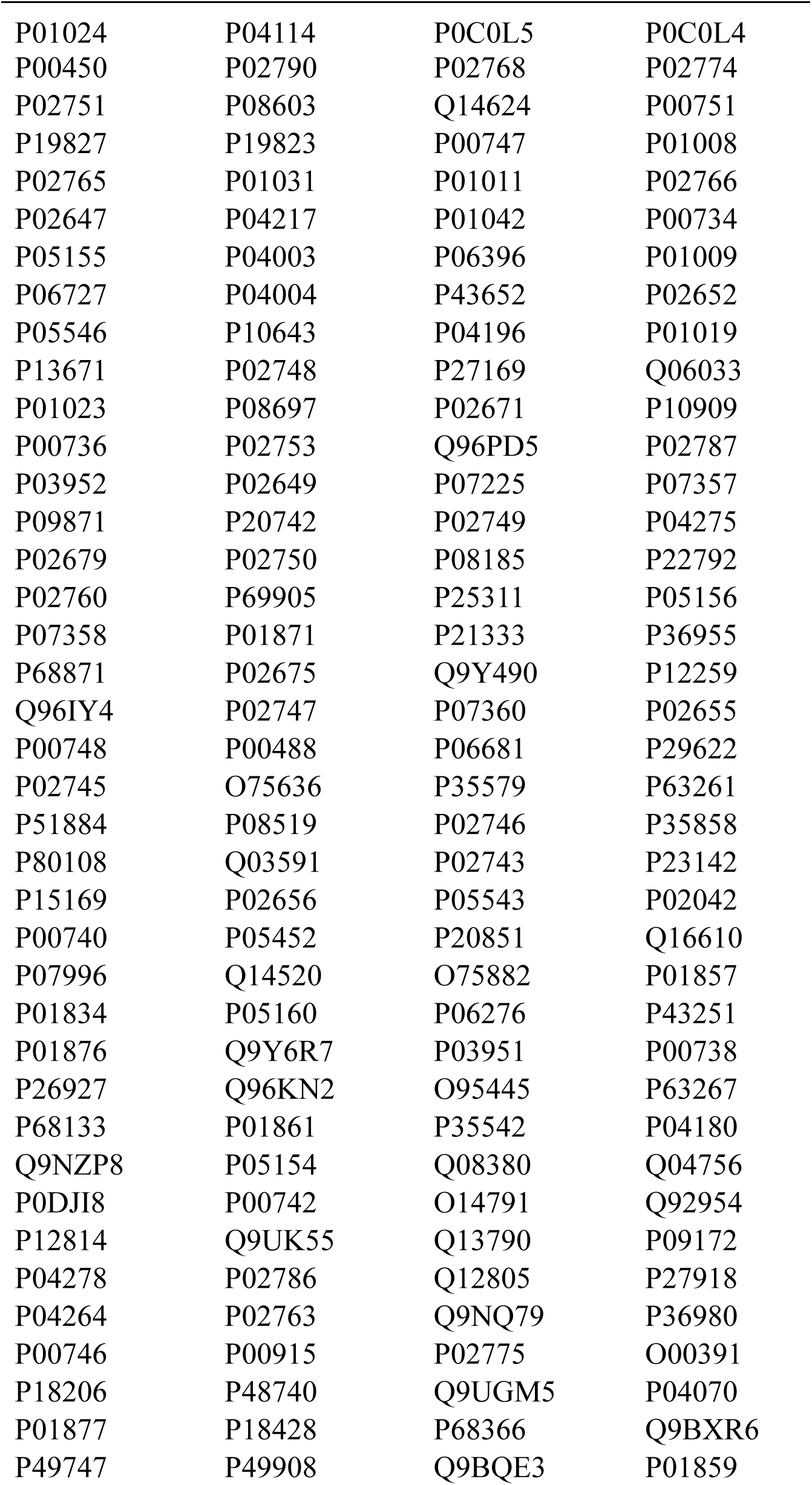
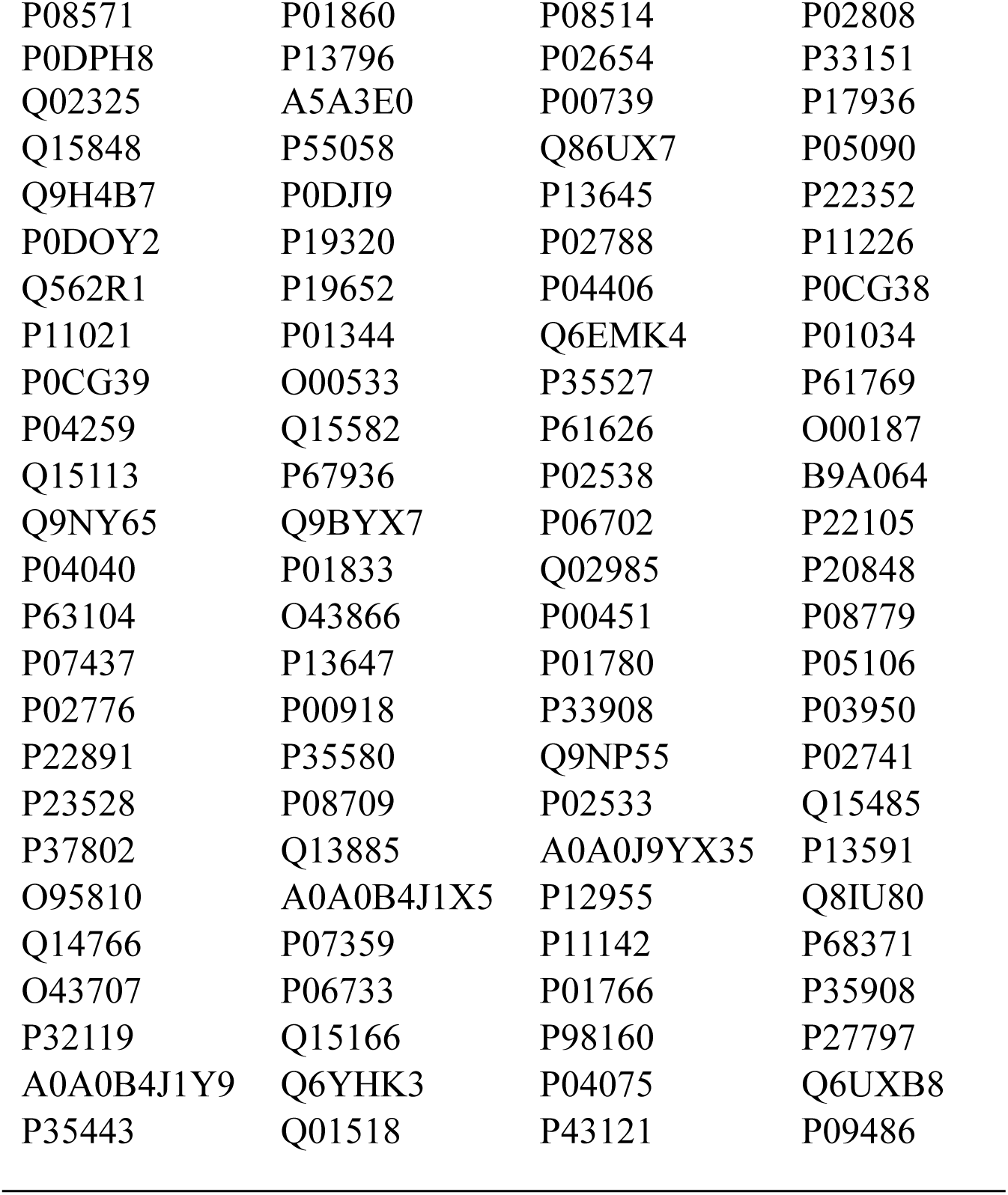
Proteomics Profiles Analyzed in this Study.

| Protein ID |  |  |  |
| --- | --- | --- | --- |
| P01024 | P04114 | P0C0L5 | P0C0L4 |
| P00450 | P02790 | P02768 | P02774 |
| P02751 | P08603 | Q14624 | P00751 |
| P19827 | P19823 | P00747 | P01008 |
| P02765 | P01031 | P01011 | P02766 |
| P02647 | P04217 | P01042 | P00734 |
| P05155 | P04003 | P06396 | P01009 |
| P06727 | P04004 | P43652 | P02652 |
| P05546 | P10643 | P04196 | P01019 |
| P13671 | P02748 | P27169 | Q06033 |
| P01023 | P08697 | P02671 | P10909 |
| P00736 | P02753 | Q96PD5 | P02787 |
| P03952 | P02649 | P07225 | P07357 |
| P09871 | P20742 | P02749 | P04275 |
| P02679 | P02750 | P08185 | P22792 |
| P02760 | P69905 | P25311 | P05156 |
| P07358 | P01871 | P21333 | P36955 |
| P68871 | P02675 | Q9Y490 | P12259 |
| Q96IY4 | P02747 | P07360 | P02655 |
| P00748 | P00488 | P06681 | P29622 |
| P02745 | O75636 | P35579 | P63261 |
| P51884 | P08519 | P02746 | P35858 |
| P80108 | Q03591 | P02743 | P23142 |
| P15169 | P02656 | P05543 | P02042 |
| P00740 | P05452 | P20851 | Q16610 |
| P07996 | Q14520 | O75882 | P01857 |
| P01834 | P05160 | P06276 | P43251 |
| P01876 | Q9Y6R7 | P03951 | P00738 |
| P26927 | Q96KN2 | O95445 | P63267 |
| P68133 | P01861 | P35542 | P04180 |
| Q9NZP8 | P05154 | Q08380 | Q04756 |
| P0DJI8 | P00742 | O14791 | Q92954 |
| P12814 | Q9UK55 | Q13790 | P09172 |
| P04278 | P02786 | Q12805 | P27918 |
| P04264 | P02763 | Q9NQ79 | P36980 |
| P00746 | P00915 | P02775 | O00391 |
| P18206 | P48740 | Q9UGM5 | P04070 |
| P01877 | P18428 | P68366 | Q9BXR6 |
| P49747 | P49908 | Q9BQE3 | P01859 |
| P08571 | P01860 | P08514 | P02808 |
| P0DPH8 | P13796 | P02654 | P33151 |
| Q02325 | A5A3E0 | P00739 | P17936 |
| Q15848 | P55058 | Q86UX7 | P05090 |
| Q9H4B7 | P0DJI9 | P13645 | P22352 |
| P0DOY2 | P19320 | P02788 | P11226 |
| Q562R1 | P19652 | P04406 | P0CG38 |
| P11021 | P01344 | Q6EMK4 | P01034 |
| P0CG39 | O00533 | P35527 | P61769 |
| P04259 | Q15582 | P61626 | O00187 |
| Q15113 | P67936 | P02538 | B9A064 |
| Q9NY65 | Q9BYX7 | P06702 | P22105 |
| P04040 | P01833 | Q02985 | P20848 |
| P63104 | O43866 | P00451 | P08779 |
| P07437 | P13647 | P01780 | P05106 |
| P02776 | P00918 | P33908 | P03950 |
| P22891 | P35580 | Q9NP55 | P02741 |
| P23528 | P08709 | P02533 | Q15485 |
| P37802 | Q13885 | A0A0J9YX35 | P13591 |
| O95810 | A0A0B4J1X5 | P12955 | Q8IU80 |
| Q14766 | P07359 | P11142 | P68371 |
| O43707 | P06733 | P01766 | P35908 |
| P32119 | Q15166 | P98160 | P27797 |
| A0A0B4J1Y9 | Q6YHK3 | P04075 | Q6UXB8 |
| P35443 | Q01518 | P43121 | P09486 |

**Sup. Table 2.**
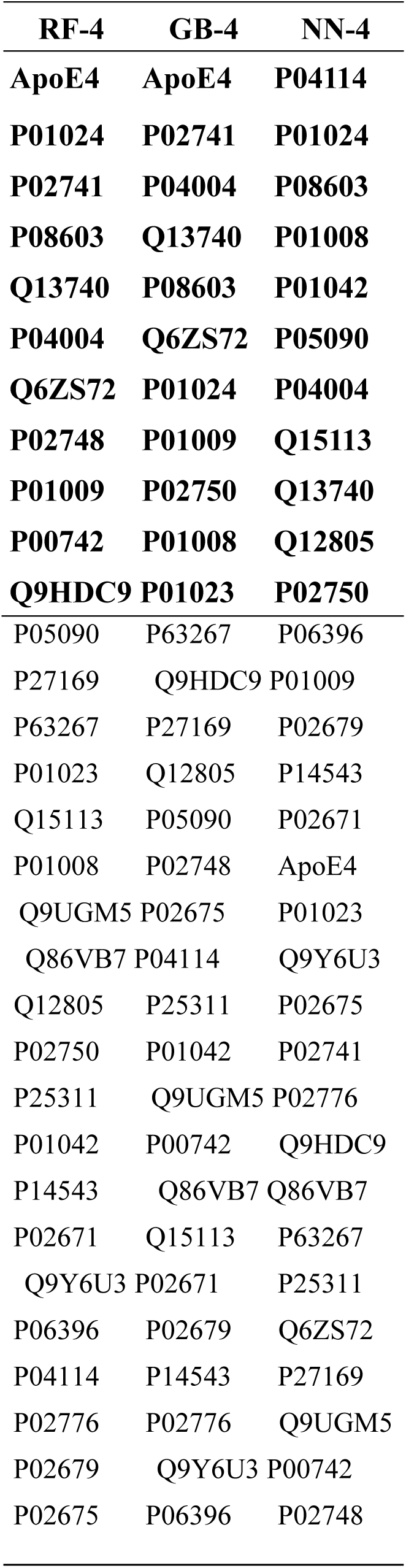
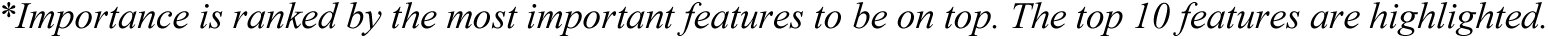
Important Features for Classification from Best Performing Model of Each Method: RF-4, GB-4, NN-4.

| **RF-4** | **GB-4** | **NN-4** |
| --- | --- | --- |
| **ApoE4** | **ApoE4** | **P04114** |
| **P01024** | **P02741** | **P01024** |
| **P02741** | **P04004** | **P08603** |
| **P08603** | **Q13740** | **P01008** |
| **Q13740** | **P08603** | **P01042** |
| **P04004** | **Q6ZS72** | **P05090** |
| **Q6ZS72** | **P01024** | **P04004** |
| **P02748** | **P01009** | **Q15113** |
| **P01009** | **P02750** | **Q13740** |
| **P00742** | **P01008** | **Q12805** |
| **Q9HDC9** | **P01023** | **P02750** |
| P05090 | P63267 | P06396 |
| P27169 | Q9HDC9 | P01009 |
| P63267 | P27169 | P02679 |
| P01023 | Q12805 | P14543 |
| Q15113 | P05090 | P02671 |
| P01008 | P02748 | ApoE4 |
| Q9UGM5 | P02675 | P01023 |
| Q86VB7 | P04114 | Q9Y6U3 |
| Q12805 | P25311 | P02675 |
| P02750 | P01042 | P02741 |
| P25311 | Q9UGM5 | P02776 |
| P01042 | P00742 | Q9HDC9 |
| P14543 | Q86VB7 | Q86VB7 |
| P02671 | Q15113 | P63267 |
| Q9Y6U3 | P02671 | P25311 |
| P06396 | P02679 | Q6ZS72 |
| P04114 | P14543 | P27169 |
| P02776 | P02776 | Q9UGM5 |
| P02679 | Q9Y6U3 | P00742 |
| P02675 | P06396 | P02748 |

**Sup. Table 3.** Differentially Expressed Proteins: Negative Vs Positive Amyloid.

| Protein | t-value | Degrees of Freedom | p-value | FDR |
| --- | --- | --- | --- | --- |
| ApoE4 | -8.307304 | 544.3554 | 0.000000000000000782 | 0.00000000000002299 |
| P01024 | 4.597768 | 668.4067 | 0.00000511 | 0.00075117 |
| P08603 | 3.691254 | 671.9609 | 0.000241 | 0.023618 |
| P02671 | -3.527214 | 454.9060 | 0.000463 | 0.0340305 |
| P04004 | 3.456623 | 671.6612 | 0.000581 | 0.0341628 |
| P01042 | 3.099841 | 671.7806 | 0.00202 | 0.09898 |
| P01009 | -3.030096 | 556.3295 | 0.00256 | 0.10752 |
| P02675 | -2.840552 | 457.5436 | 0.00470 | 0.172725 |
| P05090 | -2.746976 | 664.8847 | 0.00618 | 0.184044 |
| P25311 | -2.742384 | 659.7582 | 0.00626 | 0.184044 |
| Q6ZS72 | -2.601179 | 668.7169 | 0.00950 | 0.23618 |
| P02741 | 2.596366 | 624.1705 | 0.00964 | 0.23618 |
| P01023 | -2.530822 | 651.5998 | 0.0116 | 0.2604 |
| P06396 | -2.507754 | 665.6218 | 0.0124 | 0.2604 |
| P14543 | 2.441361 | 665.9270 | 0.0149 | 0.29204 |
| P02679 | -2.347287 | 429.7639 | 0.0194 | 0.3342316 |
| P02748 | -2.316086 | 671.9688 | 0.0209 | 0.3342316 |
| P02776 | -2.312238 | 632.6814 | 0.0211 | 0.3342316 |
| Q86VB7 | 2.302403 | 660.6239 | 0.0216 | 0.3342316 |
| P04114 | -2.226078 | 659.2533 | 0.0263 | 0.38661 |
| Q9UGM5 | 2.149521 | 670.5964 | 0.0320 | 0.410375 |
| P02750 | -2.146205 | 662.0062 | 0.0322 | 0.410375 |

PREDICTING AMYLOID POSITIVITY IN ALZHEIMER'S
| Protein | t-value | Degrees of Freedom | p-value | FDR |
| --- | --- | --- | --- | --- |
| Q9HDC9 | -2.131135 | 664.1009 | 0.0334 | 0.410375 |
| P27169 | -2.130542 | 671.3090 | 0.0335 | 0.410375 |
| Q15113 | -2.089336 | 668.6740 | 0.0371 | 0.4206462 |
| P01008 | 2.087374 | 671.8398 | 0.0372 | 0.4206462 |
| Q9Y6U3 | -2.062030 | 667.7270 | 0.0396 | 0.4312 |
| Q12805 | -2.041367 | 665.2313 | 0.0416 | 0.43512 |
| Q13740 | 2.019846 | 670.1554 | 0.0438 | 0.43512 |
| P00742 | -2.014102 | 671.8587 | 0.0444 | 0.43512 |
| P63267 | -1.981180 | 579.9825 | 0.0480 | 0.4461882 |

*\*Group-1: Negative Amyloid and Group-2: Positive Amyloid. A positive t-value (blue) indicates that the protein is upregulated in the Amyloid-negative group, while a negative t-value (red) suggests upregulation in the Amyloid-positive group.*

## References

1. Schultz, C., K.D. Tredici, and H. Braak. Neuropathology of Alzheimer’s Disease. 2004.

2. DeTure, M.A. and D.W. Dickson, The neuropathological diagnosis of Alzheimer’s disease. Mol Neurodegener, 2019. 14(1): p. 32.

3. Frisoni, G.B., et al., New landscape of the diagnosis of Alzheimer’s disease. The Lancet, 2025. 406(10510): p. 1389–1407.

4. Armstrong, R.A., The Pathogenesis of Alzheimer′s Disease: A Reevaluation of the “Amyloid Cascade Hypothesis”. International Journal of Alzheimer’s Disease, 2011. 2011(1): p. 630865.

5. Schilling, L.P., et al., Imaging Alzheimer’s disease pathophysiology with PET. Dement Neuropsychol, 2016. 10(2): p. 79–90.

6. Ewers, M., et al., CSF biomarkers for the differential diagnosis of Alzheimer’s disease: A large-scale international multicenter study. Alzheimers Dement, 2015. 11(11): p. 1306–15.

7. Leuzy, A., et al., Considerations in the clinical use of amyloid PET and CSF biomarkers for Alzheimer’s disease. Alzheimer’s & Dementia, 2025. 21(3): p. e14528.

8. Villa, C., M. Lavitrano, E. Salvatore, and R. Combi, Molecular and Imaging Biomarkers in Alzheimer’s Disease: A Focus on Recent Insights. J Pers Med, 2020. 10(3).

9. Marizzoni, M., et al., A peripheral signature of Alzheimer’s disease featuring microbiota-gut-brain axis markers. Alzheimers Res Ther, 2023. 15(1): p. 101.

10. Dhauria, M., et al., Blood-Based Biomarkers in Alzheimer’s Disease: Advancing Non-Invasive Diagnostics and Prognostics. Int J Mol Sci, 2024. 25(20).

11. Ignjatovic, V., et al., Mass Spectrometry-Based Plasma Proteomics: Considerations from Sample Collection to Achieving Translational Data. J Proteome Res, 2019. 18(12): p. 4085–4097.

12. Rehiman, S.H., et al., Proteomics as a reliable approach for discovery of blood-based Alzheimer’s disease biomarkers: A systematic review and meta-analysis. Ageing Research Reviews, 2020. 60: p. 101066.

13. Liu, Y., et al., Blood-based biomarkers of Alzheimer’s disease: Standardization and comprehensiveness. Neuroprotection, 2024. 02(04): p. 246–254.

14. Adams, C. and W. Bittremieux, A 2025 perspective on the role of machine learning for biomarker discovery in clinical proteomics. Expert Rev Proteomics, 2025: p. 1–12.

15. Climente-González, H., et al., Interpretable machine learning leverages proteomics to improve cardiovascular disease risk prediction and biomarker identification. Communications Medicine, 2025. 5(1): p. 170.

16. Carrasco-Zanini, J., et al., Proteomic prediction of diverse incident diseases: a machine learning-guided biomarker discovery study using data from a prospective cohort study. The Lancet Digital Health, 2024. 6(7): p. e470–e479.

17. Shahin-Shamsabadi, A. and J. Cappuccitti, Proteomics and machine learning: Leveraging domain knowledge for feature selection in a skeletal muscle tissue meta-analysis. Heliyon, 2024. 10(24): p. e40772.

18. Hartman, E., et al., Interpreting biologically informed neural networks for enhanced proteomic biomarker discovery and pathway analysis. Nature Communications, 2023. 14(1): p. 5359.

19. Pienaar, M.A. and K.D. Naidoo, Classification and predictive models using supervised machine learning: A conceptual review. South Afr J Crit Care, 2025. 41(1): p. e2937.

20. McHugh, C.P., M.H.S. Clement, and M. Phatak, AD Workbench: Transforming Alzheimer’s research with secure, global, and collaborative data sharing and analysis. Alzheimers Dement, 2025. 21(5): p. e70278.

21. Zhang, J., et al., Quantitative proteomics of cerebrospinal fluid from patients with Alzheimer disease. J Alzheimers Dis, 2005. 7(2): p. 125–33; discussion 173-80.

22. Wang, X., et al., Identification of methylation-regulated genes modulating microglial phagocytosis in hyperhomocysteinemia-exacerbated Alzheimer’s disease. Alzheimers Res Ther, 2023. 15(1): p. 164.

23. Gershov, D., S. Kim, N. Brot, and K.B. Elkon, C-Reactive protein binds to apoptotic cells, protects the cells from assembly of the terminal complement components, and sustains an antiinflammatory innate immune response: implications for systemic autoimmunity. J Exp Med, 2000. 192(9): p. 1353–64.

24. Irizarry, M.C., Biomarkers of Alzheimer disease in plasma. NeuroRx, 2004. 1(2): p. 226–34.

25. Hurko, O. and J.L. Ryan, Translational research in central nervous system drug discovery. NeuroRx, 2005. 2(4): p. 671–82.

26. Carter, C.J., Convergence of genes implicated in Alzheimer’s disease on the cerebral cholesterol shuttle: APP, cholesterol, lipoproteins, and atherosclerosis. Neurochem Int, 2007. 50(1): p. 12–38.

27. Deelen, J., et al., Genome-wide association study identifies a single major locus contributing to survival into old age; the APOE locus revisited. Aging Cell, 2011. 10(4): p. 686–98.

28. Haapasalo, K., et al., Complement Factor H Binds to Human Serum Apolipoprotein E and Mediates Complement Regulation on High Density Lipoprotein Particles. J Biol Chem, 2015. 290(48): p. 28977–87.

29. Calero, M., A. Rostagno, and J. Ghiso, Search for amyloid-binding proteins by affinity chromatography. Methods Mol Biol, 2012. 849: p. 213–23.

30. Forte, T.M., et al., Altered activities of anti-atherogenic enzymes LCAT, paraoxonase, and platelet-activating factor acetylhydrolase in atherosclerosis-susceptible mice. J Lipid Res, 2002. 43(3): p. 477–85.

31. Silverman, S.M., et al., C3- and CR3-dependent microglial clearance protects photoreceptors in retinitis pigmentosa. J Exp Med, 2019. 216(8): p. 1925–1943.

32. He, X.M., et al., Gln192Arg polymorphism in paraoxonase 1 gene is associated with Alzheimer disease in a Chinese Han ethnic population. Chin Med J (Engl), 2006. 119(14): p. 1204–9.

33. Kononikhin, A.S., et al., Prognosis of Alzheimer’s Disease Using Quantitative Mass Spectrometry of Human Blood Plasma Proteins and Machine Learning. Int J Mol Sci, 2022. 23(14).

34. Song, M., et al., Diagnostic Classification and Biomarker Identification of Alzheimer’s Disease with Random Forest Algorithm. Brain Sci, 2021. 11(4).

35. Ivarsson Orrelid, C., et al., Applying machine learning to high-dimensional proteomics datasets for the identification of Alzheimer’s disease biomarkers. Fluids and Barriers of the CNS, 2025. 22(1): p. 23.

36. Chernyaeva, L., et al., Reduced binding of apoE4 to complement factor H promotes amyloid-β oligomerization and neuroinflammation. EMBO Rep, 2023. 24(7): p. e56467.

37. Walker, D.G. and P.L. McGeer, Vitronectin expression in Purkinje cells in the human cerebellum. Neurosci Lett, 1998. 251(2): p. 109–12.

38. Shin, T.M., et al., Formation of soluble amyloid oligomers and amyloid fibrils by the multifunctional protein vitronectin. Mol Neurodegener, 2008. 3: p. 16.

39. Ruzha, Y., et al., Role of Vitronectin and Its Receptors in Neuronal Function and Neurodegenerative Diseases. International Journal of Molecular Sciences, 2022. 23(20): p. 12387.

40. Barba, L., et al., Specific Cerebrospinal Fluid SerpinA1 Isoform Pattern in Alzheimer’s Disease. Int J Mol Sci, 2022. 23(13).

41. Wu, T., et al., Complement C3 Is Activated in Human AD Brain and Is Required for Neurodegeneration in Mouse Models of Amyloidosis and Tauopathy. Cell Rep, 2019. 28(8): p. 2111–2123.e6.

42. Tao, Q., et al., Impact of C-Reactive Protein on Cognition and Alzheimer Disease Biomarkers in Homozygous APOE ɛ4 Carriers. Neurology, 2021. 97(12): p. e1243–e1252.

43. Hegazy, S.H., et al., C-reactive protein levels and risk of dementia-Observational and genetic studies of 111,242 individuals from the general population. Alzheimers Dement, 2022. 18(11): p. 2262–2271.

44. Liu, C.C., et al., Apolipoprotein E and Alzheimer disease: risk, mechanisms and therapy. Nat Rev Neurol, 2013. 9(2): p. 106–18.

45. Montagne, A., et al., APOE4 leads to blood–brain barrier dysfunction predicting cognitive decline. Nature, 2020. 581(7806): p. 71–76.

46. Leduc, V. and J. Poirier, Polymorphisms at the paraoxonase 1 L55M and Q192R loci affect the pathophysiology of Alzheimer’s disease: emphasis on the cholinergic system and beta-amyloid levels. Neurodegener Dis, 2008. 5(3-4): p. 225–7.

47. Seow, D.C., et al., Profile of the Paraoxonase 1 (PON1) Gene 192Q/R Polymorphism and Clinical Associations among Older Singaporean Chinese with Alzheimer’s and Mixed Dementia. Dement Geriatr Cogn Dis Extra, 2016. 6(1): p. 43–54.

48. Cervellati, C., et al., Evaluating the link between Paraoxonase-1 levels and Alzheimer’s disease development. Minerva Med, 2019. 110(3): p. 238–250.

49. Shen, Y., et al., *Integrated analyses of 5 mC*, *5hmC methylation and gene expression reveal pathology-associated AKT3 gene and potential biomarkers for Alzheimer’s disease*. J Psychiatr Res, 2024. 178: p. 367–377.

50. Ataka, T., et al., Plasma amyloid beta biomarkers predict amyloid positivity and longitudinal clinical progression in mild cognitive impairment. Alzheimers Dement (N Y), 2024. 10(4): p. e70008.

51. Blennow, K., et al., The potential clinical value of plasma biomarkers in Alzheimer’s disease. Alzheimer’s & Dementia, 2023. 19(12): p. 5805–5816.

52. Cheng, L., et al., Plasma Aβ as a biomarker for predicting Aβ-PET status in Alzheimer’s disease: a systematic review with meta-analysis. J Neurol Neurosurg Psychiatry, 2022. 93(5): p. 513–520.

53. Golovanevsky, M., C. Eickhoff, and R. Singh, Multimodal attention-based deep learning for Alzheimer’s disease diagnosis. Journal of the American Medical Informatics Association, 2022. 29(12): p. 2014–2022.

54. Zeng, X., et al., Multi-analyte proteomic analysis identifies blood-based neuroinflammation, cerebrovascular and synaptic biomarkers in preclinical Alzheimer’s disease. Molecular Neurodegeneration, 2024. 19(1): p. 68.

55. Dammer, E.B., et al., Multi-platform proteomic analysis of Alzheimer’s disease cerebrospinal fluid and plasma reveals network biomarkers associated with proteostasis and the matrisome. Alzheimers Res Ther, 2022. 14(1): p. 174.

